# Epidemiology of genital human papillomavirus infections in sequential male sex partners of young females

**DOI:** 10.1101/2023.06.19.23291543

**Authors:** Andrew W. Arthur, Mariam El-Zein, Ann N. Burchell, Pierre-Paul Tellier, Francois Coutlée, Eduardo L. Franco

**Affiliations:** Division of Cancer Epidemiology, McGill University, Montréal, Québec, H4A 3T2, Canada; Department of Family and Community Medicine and MAP Centre for Urban Health Solutions, Li Ka Shing Knowledge Institute, St. Michael’s Hospital, Unity Health Toronto, Toronto, Ontario, M5B 1W8, Canada; Department of Family Medicine, McGill University, Montréal, Québec, H3S 1Z1, Canada; Départements de Clinique de Médecine de Laboratoire et de Médecine, Services de Biologie Moléculaire et d’Infectiologie, Centre Hospitalier de l’Université de Montréal, Montréal, Québec, H2X 0C1, Canada

**Author notes:** **Correspondence:** Eduardo Franco, Division of Cancer Epidemiology, McGill University, Suite 720, 5100 Maisonneuve Blvd West, Montréal, Québec, H4A 3T2, Canada.

**Keywords:** Papillomaviridae, human papillomavirus, sexually transmitted infection, prospective cohort study, couple-based study, concordance, transmission, sexual partnerships, young adults

## Abstract

**Objectives:** Couple-based studies have considered human papillomavirus (HPV) transmission between current heterosexual partners (male↔female). Using data from young women and their sequential male partners in the HPV Infection and Transmission among Couples through Heterosexual activity (HITCH) study, we analysed HPV transmission from upstream sexual partnerships (male 1↔female) to downstream sex partners (→male 2).

**Methods:** Among 502 females enrolled in the HITCH study (2005-2011, Montréal, Canada), 42 brought one male sex partner at baseline (male 1) and another during follow-up (male 2). Female genital samples, collected at 6 visits over 24 months, and male genital samples, collected at 2 visits over 4 months, were tested for 36 HPV types (*n*=1512 detectable infections). We calculated observed/expected ratios with 95% confidence intervals (CIs) for type-specific HPV concordance between males 1 and 2. Using mixed-effects regression, we estimated odds ratios (ORs) with 95% CIs for male 2 testing positive for the same HPV type as male 1.

**Results:** Detection of the same HPV type in males 1 and 2 occurred 2.6 times (CI:1.9-3.5) more often than chance. The OR for male 2 positivity was 4.2 (CI:2.5-7.0). Adjusting for the number of times the linking female tested positive for the same HPV type attenuated the relationship between male 1 and 2 positivity, suggesting mediation.

**Conclusions:** High type-specific HPV concordance between males 1 and 2 confirms HPV’s transmissibility in chains of sequential young adult sexual partnerships. HPV positivity in an upstream partnership predicted positivity in a downstream male when the linking female partner was persistently positive.

## INTRODUCTION

Human papillomavirus (HPV) is the most prevalent sexually transmitted infection (STI), with young adults bearing the principal disease burden [1]. Infection with oncogenic HPV types is a necessary cause of cervical cancer and a component cause of other anogenital cancers in males and females [2]. Vaccination prevents HPV infection in the individual and creates population-level herd effects, whereby limiting incident infections in vaccinated individuals prevents transmission to unvaccinated individuals [3].

To date, couple-based studies have focused on HPV concordance and transmission between male and female partners in a current sexual relationship [4-12], estimating the impact of past and concurrent partners on HPV incidence in current sexual partnerships [7, 9-12]. However, to adequately characterize HPV transmission between sequential sexual partnerships, HPV genotyping data for multiple sex partners of the same individual are required. We analysed genital HPV transmission from upstream sexual partnerships to downstream sex partners (male 1↔female→male 2 transmission) in the HPV Infection and Transmission among Couples through Heterosexual activity (HITCH) Cohort Study. We characterised type-specific HPV concordance and associations between sequential male partners of the same female and identified correlates of HPV types detected in male 1 being detected in male 2.

## METHODS

### Study Design

Full details on the HITCH study are published elsewhere [13]. Briefly, in Montréal, Canada (2005-2011), we enrolled college-aged females and their male partners within their first 6 months of sexual activity together. Self-reported sex determined enrolment (gender data were not collected). During follow-up, some females brought subsequent male partners into the study, forming a female-linked partnership (male 1-female-male 2). Each participant within the linked partnership completed questionnaires. Females provided vaginal samples at baseline and 5 follow-up visits (4-, 8-, 12-, 18- and 24-months) while males provided scrotal and penile samples at two visits: baseline and, beginning in October 2006, follow-up at 4 months. Couples were asked to refrain from penetrative sex for 24 hours before sampling. Females were instructed to collect a vaginal sample using a polyester swab (diagnostic accuracy validated [14]). Nurses collected male epithelial cells using emery exfoliation, collecting separate polyester swab samples of the scrotum and penis (glans, external meatus, coronal sulcus, shaft, and foreskin) [15]. We tested samples for 36 HPV types using the Linear Array genotyping assay (Roche Molecular Systems), assessing cellularity via β-globin DNA coamplification [16]. Noting a high degree of type-specific HPV co-detection between scrotal and penile samples, we increasingly combined male genital samples prior to genotyping over time.

The Institutional Review Boards of McGill and Concordia Universities, and the Centre Hospitalier de l’Université de Montreal approved the HITCH study; all participants provided written informed consent.

### Statistical Analyses

We treated female-linked partnerships as the units of observation and detectable HPV infections as the units of analysis. We used Cohen’s kappa to assess the degree of type-specific HPV positivity agreement between uncombined scrotal and penile samples at each visit, as well as between combined male genital samples at baseline and follow-up. Kappa values 0.41-0.60 indicated moderate, 0.61-0.80 substantial, and 0.81-1.00 almost perfect agreement [17]. We combined positivity for each male into a single measure per HPV type (i.e., HPV type *x* positivity at either/both genital site(s) at either/both visit(s)).

Analyses were performed for any HPV type and by Alphapapillomavirus subgenera. The latter analyses grouped HPV types by tissue tropism, oncogenicity and phylogenetic relatedness, generating groups of biologically and clinically comparable types. Subgenus 1 includes low oncogenic risk mucosal HPVs 6, 11, 40, 42, 44, and 54; subgenus 2 includes high oncogenic risk mucosal HPVs 16, 18, 26, 31, 33, 35, 39, 45, 51, 52, 53, 56, 58, 59, 66, 67, 68, 69, 70, 73 and 82; and subgenus 3 includes commensal mucocutaneous HPVs 61, 62, 71, 72, 81, 83, 84 and 89 [18, 19]. The ability to detect multiple HPV genotypes in the same genital sample can generate intraparticipant correlation in analyses of grouped HPV types [20]; we used statistical approaches that account for data clustering (detailed below).

Assuming HPV positivity is independent for males 1 and 2, their expected concordance is the product of infection prevalence, divided by total detectable infections. We calculated observed/expected (O/E) ratios for type-specific HPV concordance between males 1 and 2 with percentile-based 95% confidence intervals (CIs). Our 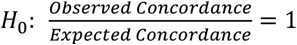 implies O/E ratio divisor/dividend correlation. Bootstrap CIs maintain nominal coverage when dividends and divisors are highly correlated [21]. Bootstraps for overall and subgenera-specific analyses resampled data by linked-partnership clusters.

We used mixed-effects logistic regression models with random intercepts by linked partnership and exchangeable correlation structure to estimate odds ratios (ORs) with 95% CIs for male 2 testing positive (outcome) for an HPV type detected in male 1 (exposure) [20]. Selecting covariates based on previous investigations of HPV transmission and natural history in the HITCH study [12, 22], we adjusted/stratified our models by covariates pertaining to HPV positivity: number of times the female partner tested positive up to male 2 enrolment, and time between the last available male 1 sample and baseline male 2 sample (< vs. ≥median). We also adjusted/stratified by sexual behaviour covariates pertaining to the downstream partnership: male 2 concurrent partners; instances of vaginal sex (< vs. ≥median); and condom use frequency during vaginal sex (≤ vs. >75%). Strata cutoffs were defined to optimize the equivalency of detectable infections between strata.

We performed additional analyses to explore the impact of differential male detection opportunities and female cell deposition on our odds estimates. For the former, we adjusted for the sum of genital samples provided by males 1 and 2. For the latter, given previous findings of male cells in vaginal specimens of HITCH-enrolled females, we adjusted and stratified (≤ vs. >3 days) by the time since vaginal sex at male 2 baseline [23]. Statistical analyses were conducted using Stata SE 17.

## RESULTS

Of 502 enrolled females, 42 brought a second male partner post-baseline, resulting in an observational sample of 42 female-linked partnerships (42 females, 82 males), and an analytical sample of 1,512 detectable HPV infections (Figure 1). The median time between the last available male 1 sample and the baseline male 2 sample was 10.2 months (interquartile range: 6.4-18.4 months; Figure S1 presents the distribution of time elapsed). Demographic and lifestyle covariates were similar between partners (Table 1). Males were, on average, older at baseline and had more lifetime vaginal sex partners compared to females. Male 2 was generally older and had more past vaginal sex partners compared to male 1. Table S1 shows moderate to strong agreement between uncombined male genital samples (kappa: 0.47-1) and between combined samples at both male visits (0.55-0.75), which justified combining baseline and follow-up penile and scrotal results.

**Table 1.**
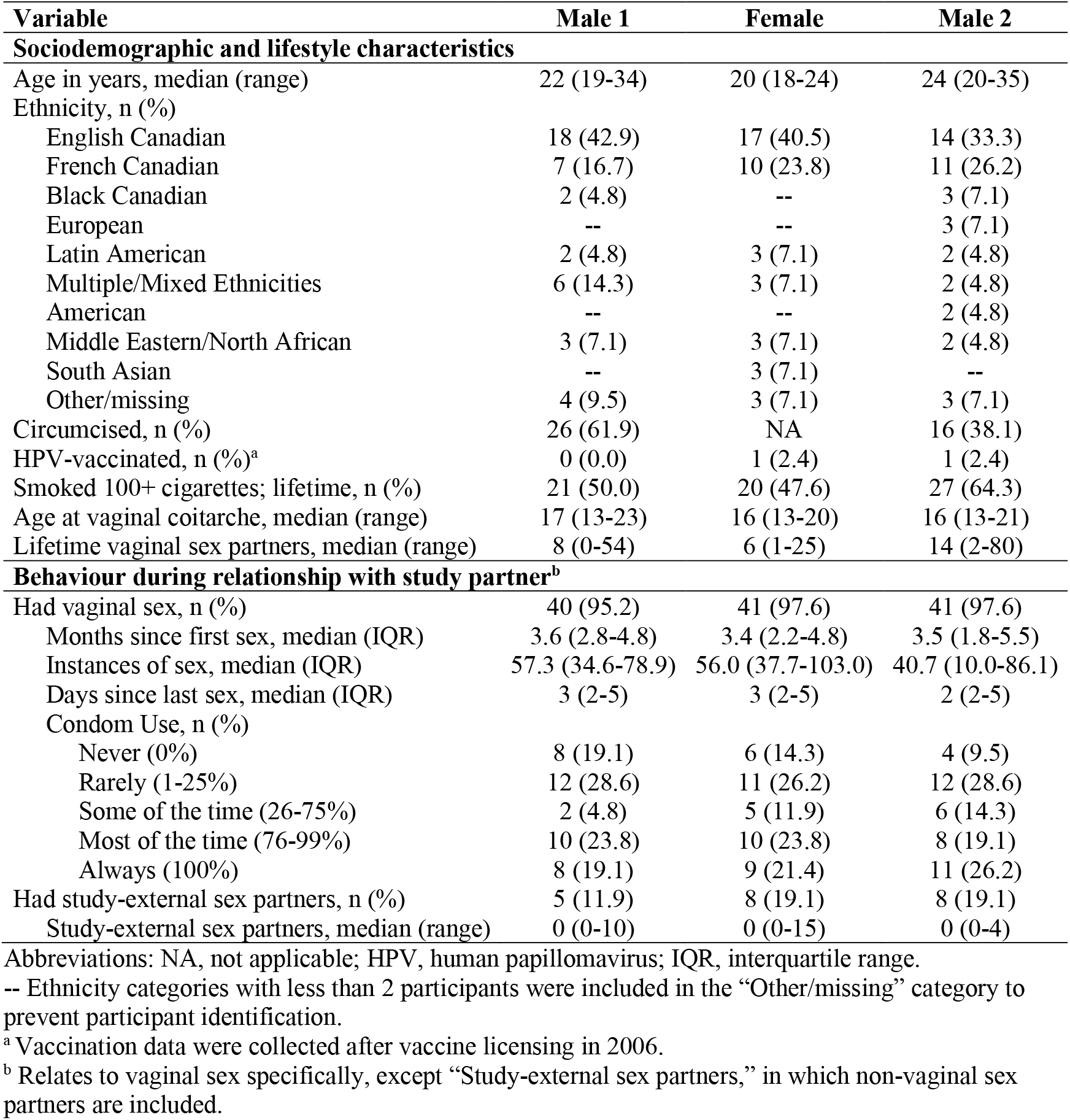
Self-reported baseline characteristics of linked partnerships in the HITCH study.

**Figure 1.**
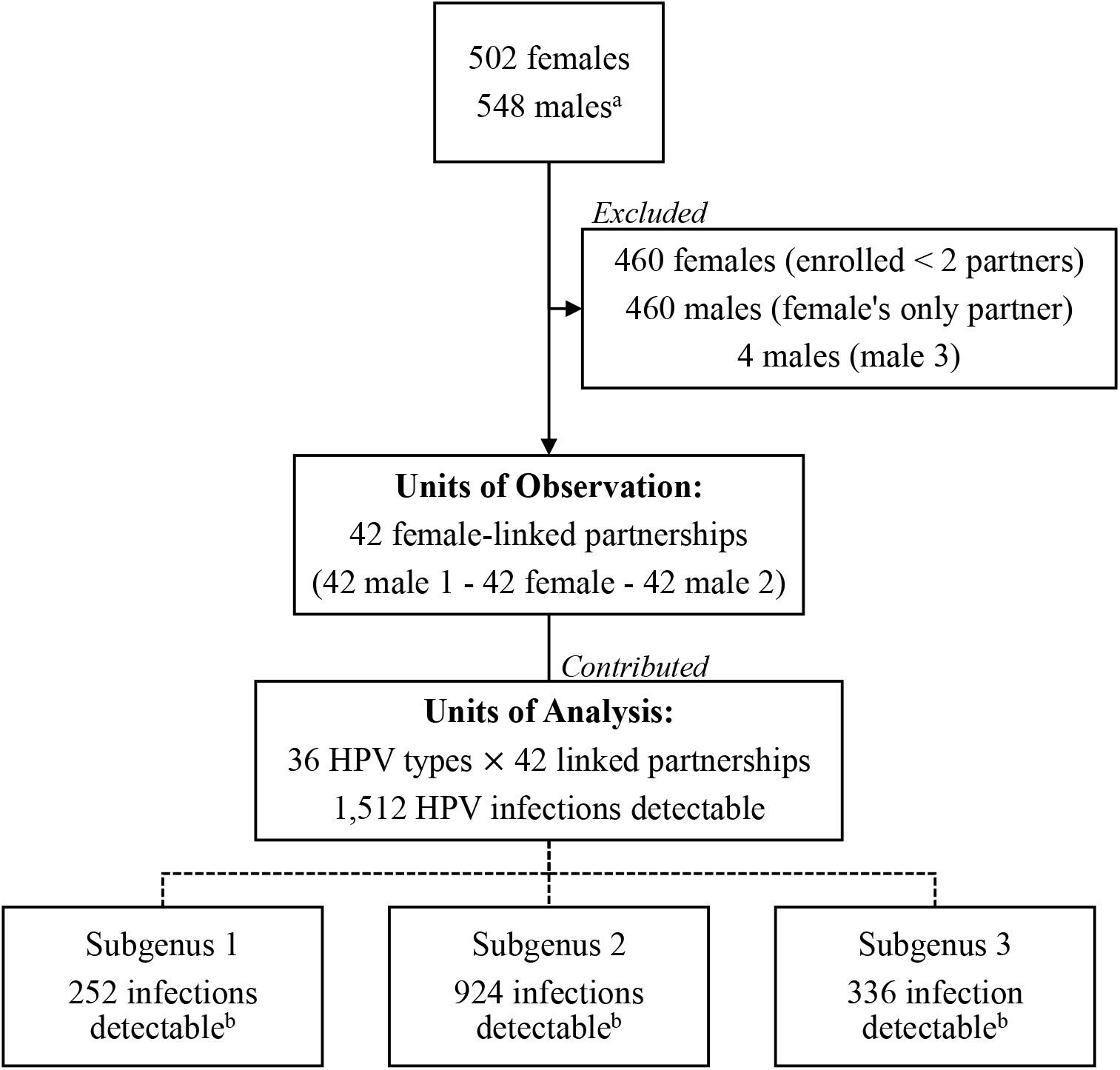
Sample selection in the HITCH study and analytic framework. → Indicates changes in observational sample size. — Indicates data transformation from observational to analysis sample. ---- Indicates breakdown of analysis sample into Alphapapillomavirus subgenera. ^a^ HITCH enrolled 548 couples. 42 women enrolled subsequent male partner(s) at follow-up. ^b^ Subgenus 1 includes low oncogenic risk mucosal HPVs 6, 11, 40, 42, 44, and 54; subgenus 2 includes high oncogenic risk mucosal HPVs 16, 18, 26, 31, 33, 35, 39, 45, 51, 52, 53, 56, 58, 59, 66, 67, 68, 69, 70, 73 and 82; and subgenus 3 includes commensal mucocutaneous HPVs 61, 62, 71, 72, 81, 83, 84 and 89 [18, 19].

Overall, concordance for any HPV infection between males 1 and 2 was observed 2.6, CI:1.9-3.5 times more often than expected (Table 2). About 24% (29/120) of HPV infections detected in male 1 were also detected in male 2. Figure S2 shows the expected type-specific concordance distribution.

**Table 2.** Type-specific HPV concordance between male 1 and male 2, overall and by subgenus.^a^

| Alphapapillomavirus | HPV Type | Positivity Male 1/Male 2 |  |  |  | Expected Concordance | Observed/Expected Concordance (95% CI) |
| --- | --- | --- | --- | --- | --- | --- | --- |
|  |  | -/- | -/+ | +/- | +/+ |  |  |
| All subgenera | Any | 1,283 | 109 | 91 | 29 | 10.95 | 2.6 (1.9-3.5) |
| Subgenus 1 | 6 | 29 | 6 | 5 | 2 | 1.33 | 1.5 (0.0-4.0) |
|  | 11 | 42 | 0 | 0 | 0 | 0 | NCE |
|  | 40 | 37 | 3 | 2 | 0 | 0.14 | 0 |
|  | 42 | 29 | 4 | 6 | 3 | 1.5 | 2.0 (0.0-4.2) |
|  | 44 | 39 | 2 | 1 | 0 | 0.05 | 0 |
|  | 54 | 35 | 3 | 3 | 1 | 0.38 | 2.6 (0.0-10.5) |
|  | Any | 211 | 18 | 17 | 6 | 2.19 | 2.7 (1.0-4.2) |
| Subgenus 2 | 16 | 27 | 5 | 4 | 6 | 2.62 | 2.3 (1.3-4.0) |
|  | 18 | 36 | 4 | 2 | 0 | 0.19 | 0 |
|  | 26 | 42 | 0 | 0 | 0 | 0 | NCE |
|  | 31 | 36 | 4 | 2 | 0 | 0.19 | 0 |
|  | 33 | 41 | 1 | 0 | 0 | 0 | NCE |
|  | 34 | 42 | 0 | 0 | 0 | 0 | NCE |
|  | 35 | 41 | 1 | 0 | 0 | 0 | NCE |
|  | 39 | 32 | 4 | 5 | 1 | 0.71 | 1.4 (0.0-4.7) |
|  | 45 | 39 | 2 | 1 | 0 | 0.05 | 0 |
|  | 51 | 22 | 10 | 8 | 2 | 2.86 | 0.7 (0.0-1.7) |
|  | 52 | 39 | 1 | 2 | 0 | 0.05 | 0 |
|  | 53 | 26 | 8 | 6 | 2 | 1.9 | 1.1 (0.0-2.4) |
|  | 56 | 29 | 8 | 4 | 1 | 1.07 | 0.9 (0.0-3.4) |
|  | 58 | 38 | 1 | 2 | 1 | 0.14 | 7.0 (0.0-21.0) |
|  | 59 | 34 | 3 | 4 | 1 | 0.48 | 2.1 (0.0-8.4) |
|  | 66 | 34 | 3 | 4 | 1 | 0.48 | 2.1 (0.0-9.3) |
|  | 67 | 32 | 6 | 4 | 0 | 0.57 | 0 |
|  | 68 | 40 | 1 | 1 | 0 | 0.02 | 0 |
|  | 69 | 42 | 0 | 0 | 0 | 0 | NCE |
|  | 70 | 39 | 2 | 1 | 0 | 0.05 | 0 |
|  | 73 | 32 | 5 | 4 | 1 | 0.71 | 1.4 (0.0- 4.7) |
|  | 82 | 36 | 3 | 3 | 0 | 0.21 | 0 |
|  | Any | 779 | 72 | 57 | 16 | 6.95 | 2.3 (1.4-3.3) |
| Subgenus 3 | 61 | 37 | 2 | 3 | 0 | 0.14 | 0 |
|  | 62 | 34 | 6 | 2 | 0 | 0.29 | 0 |
|  | 71 | 41 | 1 | 0 | 0 | 0 | NCE |
|  | 72 | 42 | 0 | 0 | 0 | 0 | NCE |
|  | 81 | 40 | 1 | 1 | 0 | 0.02 | 0 |
|  | 83 | 41 | 1 | 0 | 0 | 0 | NCE |
|  | 84 | 28 | 3 | 7 | 4 | 1.83 | 2.2 (0.8-4.2) |
|  | 89 | 30 | 5 | 4 | 3 | 1.33 | 2.3 (0.0-5.0) |
|  | Any | 293 | 19 | 17 | 7 | 1.86 | 3.8 (1.7-7.0) |

Abbreviations: HPV, human papillomavirus; CI, confidence interval; NCE, no concordance expected (inestimable).
<sup>a</sup> Subgenus 1 includes low oncogenic risk mucosal HPV types, subgenus 2 includes high oncogenic risk mucosal HPV types, and subgenus 3 includes commensal mucocutaneous HPV types [18, 19].

As shown in Table 3, if male 2 entered a sexual relationship with a female whose previous partner was positive for HPV type *x*, the odds of male 2 testing positive for that type increased 4.2-fold, CI:2.5-7.0. However, after adjusting for instances of female positivity up to male 2 enrolment, the OR was attenuated (1.4, CI:0.7-2.6).

**Table 3.** Crude, adjusted, and stratified odds ratios (95% confidence intervals) for the association of type-specific HPV positivity between male 1 and 2, overall and by subgenus.^a^

| Type of Analysis | Covariate Stratum | Overall | Subgenus 1 | Subgenus 2 | Subgenus 3 |
| --- | --- | --- | --- | --- | --- |
| Crude | -- | 4.2 (2.5-7.0) | 4.3 (1.3-14.4) | 3.4 (1.8-6.4) | 9.9 (2.8-35.4) |
| **HPV Positivity-Related Covariates** |  |  |  |  |  |
| Adjusted for instances of female positivity<sup>b</sup> | -- | 1.4 (0.7-2.6) | 0.2 (0.0-2.0) | 1.6 (0.7-3.3) | 1.0 (0.2-5.7) |
| Stratified by time between male 1 last available, and male 2 baseline samples | <10.2 months | 5.8 (2.9-11.7) | 7.5 (1.0-54.0) | 5.7 (2.4-13.6) | 6.6 (1.0-41.4) |
|  | ≥10.2 months | 2.9 (1.3-6.2) | 3.1 (0.6-15.8) | 1.7 (0.6-4.9) | 13.3 (2.3-76.7) |
| **Sexual Behaviour-Related Covariates** |  |  |  |  |  |
| Adjusted for number of male 2 concurrent partners | -- | 4.3 (2.6-7.2) | 4.5 (1.3-15.5) | 3.5 (1.8-6.6) | 10.2 (2.8-37.0) |
| Stratified by instances of sex<sup>c</sup> | < 40.7 instances | 2.5 (1.1-5.7) | NC | 2.3 (0.9-6.3) | 5.3 (1.1-26.5) |
|  | ≥ 40.7 instances | 6.4 (3.2-12.8) | 11.3 (1.7-73.2) | 4.7 (2.0-11.4) | 29.6 (2.9-297.7) |
| Stratified by frequency of condom use<sup>c</sup> | ≤ 75% of the time | 4.6 (2.4-8.8) | 4.5 (1.1-18.4) | 6.1 (2.6-14.2) | 2.2 (0.4-13.9) |
|  | > 75% of the time | 3.8 (1.7-8.7) | 2.7 (0.2-29.8) | 1.5 (0.5-4.6) | 55.7 (5.9-527.1) |
| **Additional Analyses** |  |  |  |  |  |
| Adjusted for sum of male 1 & 2 samples (detection opportunity) | -- | 4.2 (2.5-7.0) | 4.6 (1.4-15.4) | 3.3 (1.7-6.4) | 9.7 (2.7-34.7) |
| Adjusted for recency of sex, continuous (female deposition)<sup>c</sup> | -- | 4.3 (2.6-7.1) | 4.4 (1.3-14.9) | 3.4 (1.8-6.6) | 10.8 (2.9-40.1) |
| Stratified by recency of sex, binary (female deposition)<sup>c</sup> | ≤ 3 days prior | 6.0 (3.3-11.0) | 5.2 (1.4-20.2) | 4.4 (2.1-9.5) | 17.0 (3.6-81.2) |
|  | > 3 days prior | 1.3 (0.4-4.6) | NC | 1.7 (0.5-6.6) | NC |

Abbreviations: HPV, human papillomavirus; NC, no concordance.
-- indicates not applicable.
<sup>a</sup> Subgenus 1 includes low oncogenic risk mucosal HPVs 6, 11, 40, 42, 44, and 54; subgenus 2 includes high oncogenic risk mucosal HPVs 16, 18, 26, 31, 33, 35, 39, 45, 51, 52, 53, 56, 58, 59, 66, 67, 68, 69, 70, 73 and 82; and subgenus 3 includes commensal mucocutaneous HPVs 61, 62, 71, 72, 81, 83, 84 and 89 [18, 19].
<sup>b</sup> Up to male 2 enrolment.
<sup>c</sup> Relates to vaginal sex in the upstream partnership, as reported by male 2 at baseline; 1 linked partnership was excluded (no vaginal sex). 1 additional linked partnership was excluded from analyses stratified by instances of sex (missing data).

The following covariates lowered the odds that male 2 would test positive for the same HPV type as male 1: time gaps between male 1 and 2 sampling ≥10.2 months (2.9, CI:1.3-6.2), <40.7 instances of vaginal sex in the downstream partnership (2.5, CI:1.1-5.7), and condom use >75% of the time in the downstream partnership (3.8, CI:1.7-8.7). The odds of male 2 positivity were not strongly impacted by male 2 concurrent sex partner count (4.3, CI:2.6-7.2). Concerning analyses by subgenera, the relationship between HPV detection in males 1 and 2 was often unique for subgenus 3. While O/E ratios for subgenera 1 and 2 were similar to the overall estimate, that of subgenus 3 was larger (3.8, CI:1.7-7.0). The OR for male 2 testing positive for the same HPV type as male 1 was also larger for subgenus 3 (9.9, CI:2.8-35.4), while ORs for subgenera 1 and 2 were similar to the overall estimate. Additionally, for subgenus 3, the OR was higher when the time gap between male 1 and 2 sampling was ≥ 10.2 months, and when the downstream partnership was using condoms ≤ 75% of the time. Both covariates had the opposite influence on the OR estimate for subgenera 1 and 2.

Adjusting for the total number of genital samples provided by males 1 and 2 (i.e., male detection opportunities) did not change the overall OR. The number of days since the downstream partnerships’ most recent vaginal sex encounter had a minimal impact on the OR (4.3, CI:2.6-7.1). However, treated as a binary variable, the OR differed between downstream partnerships who had vaginal sex ≤ 3 days ago (6.0, CI:3.3-11.0) vs. > 3 days ago (1.3, CI:0.4-4.6).

## DISCUSSION

In this study of sequential young adult heterosexual relationships, we found evidence of HPV infections originating in upstream relationships being transmitted to downstream partners via a linking partner (male 1↔female→male 2). Detection of the same HPV type in males 1 and 2 was observed 2.6 times more often than chance, indicating that HPV positivity in a female’s sequential sex partners is not independent. The 4.2-fold higher odds that male 2 would test positive for an HPV type detected in male 1 were attenuated by the female partner’s positivity for the same type, suggesting mediation via the linking female partner. When we adjusted for the number of female detections, each additional instance of type-specific HPV detection in the female partner increased the odds of male 2 positivity 2.2-fold, CI:1.8-2.7, on average. Hence, infection persistence in the linking female appears to be relevant to whether a particular HPV type becomes detectable in the downstream male.

To our knowledge, this is the first couple-based analysis of HPV transmission across sequential heterosexual relationships. Notwithstanding, our results are unsurprising in the context of couple-based analyses of current sexual partnerships. Type-specific HPV concordance between current partners is consistently higher than expected [4-6, 8, 10, 11]; a current partner’s positivity is a strong determinant for an uninfected partner’s incident infection via type-specific HPV transmission [7-12].

Gaps ≥ 10.2 months between testing males 1 and 2 for HPV considerably lowered the odds of male 2 testing positive. This variable may be a crude proxy of the time elapsed between downstream and upstream partnerships, which is a risk factor for STI diagnosis in downstream partners [24]. We recently estimated that genital infections persisted, on average, 10.3-14.8 months in HITCH study-enrolled women [22], which corroborates a role for female infection persistence in female→male 2 transmission. Presumably, infection clearance or latency in the linking partner before initiation of the downstream partnership decreases the odds of transmission to a subsequent partner.

Several sexual behaviours in the downstream partnership were relevant to the odds of male 2 testing positive for the same HPV type as male 1. We observed, in accordance with couple-based transmission studies, an increased risk of infection with increasing instances of vaginal sex [7, 10, 12] and decreasing frequency of condom use [7, 12]. Having concurrent partners had little impact on male 2 type-specific HPV concordance with male 1. This is expected; male 2 infection via a concurrent partner is a function of his concurrent partner’s infection status, which should be independent of male 1 type-specific HPV positivity.

Compared to subgenera 1 and 2, subgenus 3 HPV types had large O/E ratios and crude ORs. Our estimates of similar infection persistence across subgenera in the HITCH cohort [22] cannot explain higher odds of male 2 positivity with longer timelapses between testing male partners. Subgenus 3 HPV types also reversed the typically odds-lowering impact of more-frequent condom use observed for subgenera 1 and 2. Their mucocutaneous tissue tropism may have allowed subgenus 3 HPV types to spread via sexual contact between tissue not covered by condoms (e.g., pubic hair follicles) [19].

The key strengths of our study and analysis are enrolment of linked partnerships and longitudinal observation of the female partner. We acknowledge four important limitations. Firstly, while partners were asked to refrain from sex 24 hours before sampling, we have estimated that 14.1% of detections in males and females in the HITCH study are attributable to deposition [25]. Recency of vaginal sex, considered on a continuous scale, minimally impacted the OR. However, we have detected Y chromosome DNA in female genital samples up to 3 days after vaginal sex in this cohort [23], and the odds of male 2 testing positive were insignificant when downstream partnerships’ most-recent sex was ≤3 days ago. While we expect opposite-sex cells to dissipate faster from male genitalia during routine washing, cleaning the mucosal glans penis may be less common in uncircumcised males (60% of male 2) [26]. Secondly, combining male HPV DNA positivity into one measurement may have introduced misclassification. However, adjusting for the number of times males were tested did not change OR estimates, suggesting that combining genital sample results did not strongly influence our estimates despite differential detection opportunities. Thirdly, we ascertained sexual behaviour covariates via male 2 self-report. However, inter-partner reporting discrepancies were minimal for the upstream partnership, and we expect the same to hold true for the downstream partnership. Finally, the small number of linked partnerships and HPV types in our mixed effects models, as well as the low prevalence of the outcome (9%, overall) may have biased our fixed effects and variance-covariance estimates [27]. Future studies would benefit from larger numbers of linked partnerships and the assessment of female↔male→female transmission directionalities.

Our findings may have implications for HPV-related cancer prevention strategies. Unvaccinated females and males gain protection from HPV infection via herd effects when vaccinated individuals do not contract, and subsequently do not transmit the virus [3]. According to our findings, had male 2 not entered a partnership with a persistently positive female whose previous partner tested positive, the odds that male 2 would test positive for the same infection would diminish. Vaccinating males may therefore prevent infection in both current unvaccinated sex partners (“direct” herd effects) *and* downstream unvaccinated sexual connections (“indirect” herd effects). While vaccinating females alone probably has “indirect” herd effects, males tend to have more multiple/recent sex partners compared to females [28], and herd effects are augmented when vaccine coverage is high in groups that are more sexually active [3].

Our results provide evidence that HPV is transmissible in a chain of sequential heterosexual young adult partnerships. Positivity in an upstream partnership is a strong predictor of type-specific HPV positivity in a downstream male partner when the linking female partner is persistently positive for that type. These estimates suggest that vaccinating upstream male partners may exert “indirect” herd effects on downstream sexual connections.

## Supporting information

STROBE Cohort

## Data Availability

HITCH participant consent forms specified that data would be published in aggregate form and that individual-level data would only be available to study investigators. To access individual-level data, please contact Eduardo Franco at. Analytical codes and a data dictionary are available on the McGill University Dataverse.

https://doi.org/10.5683/SP3/KNNHSR

## NOTES

### Disclaimer

The funders played no role in study design, data collection/analysis, preparation of the manuscript, or the decision to submit it for publication.

### Ethics Approval

HITCH complies with all national/international regulations regarding research with human data and materials, including the Declaration of Helsinki. The study was conducted per the principles and articles stipulated by the Tri-Council Policy Statement: Ethical Conduct for Research Involving Humans. Ethical approval was obtained from the institutional review boards at McGill University, Concordia University, and the Centre Hospitalier de l’Université de Montréal. Ethics renewal approval is requested annually from McGill University (study number A09-M77-04A). All subjects provided written informed consent for study participation.

## Acknowledgements

We thank the volunteering participants and employees of the HITCH cohort study, as well as the staff of the Student Health Services Clinics at McGill and Concordia universities. We also thank Dr. Talía Malagón for her guidance in selecting and operationalizing statistical analyses. Portions of this manuscript were presented at the 23^rd^ Annual McGill Biomedical Graduate Conference, March 2023, Montréal, QC, Canada; at a McGill University, Department of Oncology seminar, June 2023, Montréal, QC, Canada; at the McGill University Celebration of Research and Training in Oncology, June 2023, Montréal, QC, Canada; and at the Canadian Society for Epidemiology and Biostatistics Conference, June 2023, Halifax, NS, Canada.

## Access to Data

HITCH participant consent forms specified that data would be published in aggregate form and that individual-level data would only be available to study investigators. To access individual-level data, please contact Eduardo Franco at. The protocol for the HITCH cohort study has been published [13]. Analytical codes and a data dictionary are available on the McGill University Dataverse [29].

## Author Contributions

Conceptualization: A.N.B. (equal, HITCH), E.L.F. (lead); Funding Acquisition: A.N.B. (lead, HITCH), E.L.F. (lead, HITCH); Investigation: A.N.B. (lead, HITCH), P.P.T. (equal, HITCH), F.C. (equal, HITCH); Project Administration: A.N.B. (lead, HITCH), P.P.T. (equal, HITCH), E.L.F. (equal, HITCH); Resources: P.P.T. (equal, HITCH), F.C. (equal, HITCH); Data curation: M.Z. (equal, HITCH), A.N.B. (equal, HITCH); Supervision: M.Z. (equal), A.N.B. (supporting), E.L.F. (lead); Methodology: A.W.A. (lead), M.Z. (equal), E.L.F. (equal); Formal Analysis: A.W.A. (lead); Visualization: A.W.A. (lead); Validation: A.W.A. (lead), F.C. (lead, HITCH); Writing – original draft: A.W.A. (lead); Writing – review and editing: M.Z. (lead), A.N.B. (equal), P.P.T. (equal), F.C. (equal), E.L.F. (equal).

