## Supplemental Material for "Epidemiology of genital human papillomavirus infections in sequential male sex partners of young females"

**Table S1.** Agreement between sample collection sites and visits in males for type-specific HPV positivity, overall and by subgenus.[a]

| | **Penile and Scrotal Samples at Baseline** | | | | | **Penile and Scrotal Samples at Follow-up** | | | | | **Baseline and Follow-up: Combined Genital Site(s)** | | | | |
|---|---|---|---|---|---|---|---|---|---|---|---|---|---|---|---|
| | **--** | **-+** | **+-** | **++** | **Kappa[b]** | **--** | **-+** | **+-** | **++** | **Kappa[b]** | **--** | **-+** | **+-** | **++** | **Kappa[b]** |
| **Male 1** | | | | | | | | | | | | | | | |
| **Overall** | 591 | 5 | 15 | 37 | 0.77 | 712 | 6 | 18 | 20 | 0.61 | 1154 | 18 | 27 | 61 | 0.71 |
| **Subgenus 1** | 98 | 1 | 3 | 6 | 0.73 | 116 | 1 | 5 | 4 | 0.55 | 190 | 3 | 6 | 11 | 0.69 |
| **Subgenus 2** | 362 | 1 | 11 | 22 | 0.77 | 435 | 4 | 13 | 10 | 0.52 | 706 | 12 | 15 | 37 | 0.71 |
| **Subgenus 3** | 131 | 3 | 1 | 9 | 0.80 | 161 | 1 | 0 | 6 | 0.92 | 258 | 3 | 6 | 13 | 0.73 |
| **Male 2** | | | | | | | | | | | | | | | |
| **Overall** | 497 | 4 | 22 | 17 | 0.54 | 168 | 1 | 2 | 9 | 0.85 | 999 | 37 | 26 | 54 | 0.60 |
| **Subgenus 1** | 82 | 0 | 4 | 4 | 0.65 | 29 | 0 | 0 | 1 | 1.00 | 164 | 4 | 4 | 14 | 0.75 |
| **Subgenus 2** | 303 | 3 | 15 | 9 | 0.47 | 100 | 1 | 2 | 7 | 0.81 | 609 | 25 | 17 | 31 | 0.56 |
| **Subgenus 3** | 112 | 1 | 3 | 4 | 0.65 | 39 | 0 | 0 | 1 | 1.00 | 226 | 8 | 5 | 9 | 0.55 |

Abbreviation: HPV, human papillomavirus.

[a] Subgenus 1 includes low oncogenic risk mucosal HPVs 6, 11, 40, 42, 44, and 54; subgenus 2 includes high oncogenic risk mucosal HPVs 16, 18, 26, 31, 33, 35, 39, 45, 51, 52, 53, 56, 58, 59, 66, 67, 68, 69, 70, 73 and 82; and subgenus 3 includes commensal mucocutaneous HPVs 61, 62, 71, 72, 81, 83, 84 and 89 [18, 19].

[b] Kappa values 0.41-0.60 indicate moderate, 0.61-0.80 substantial, and 0.81-1.00 almost perfect agreement [17].

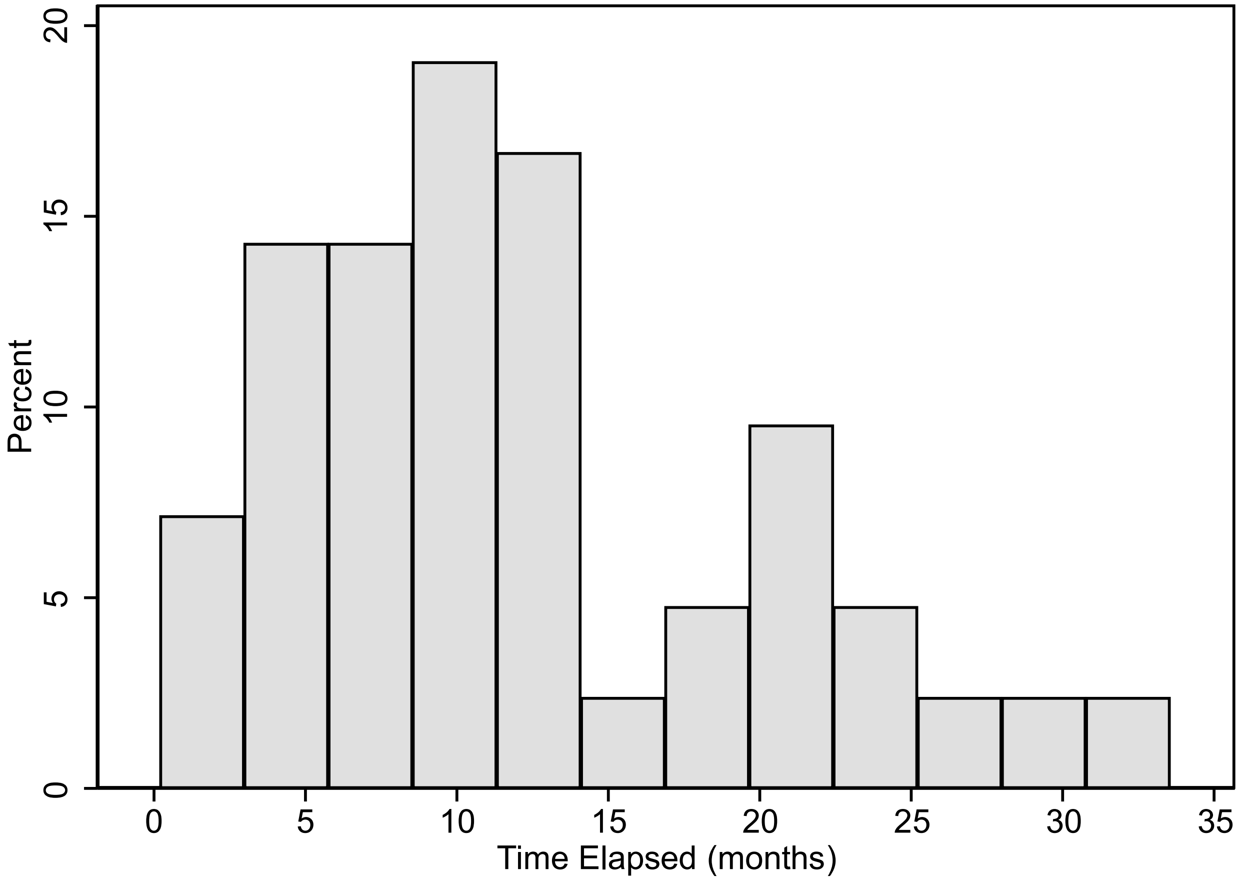


**Figure S1.** Distribution of time elapsed between male 1 last available sample and male 2 baseline sample within linked partnerships.

**Figure S1 Legend:** While all male 2 provided a baseline genital sample, male 1 follow-up visits did not begin until October 2006. As such, we measured the time between available male 1 and male 2 genital samples based on the final sample male 1 provided. When male 1 provided a follow-up sample, we measured the time between male 1 follow-up and male 2 baseline; otherwise, we measured the time between male 1 baseline and male 2 baseline.

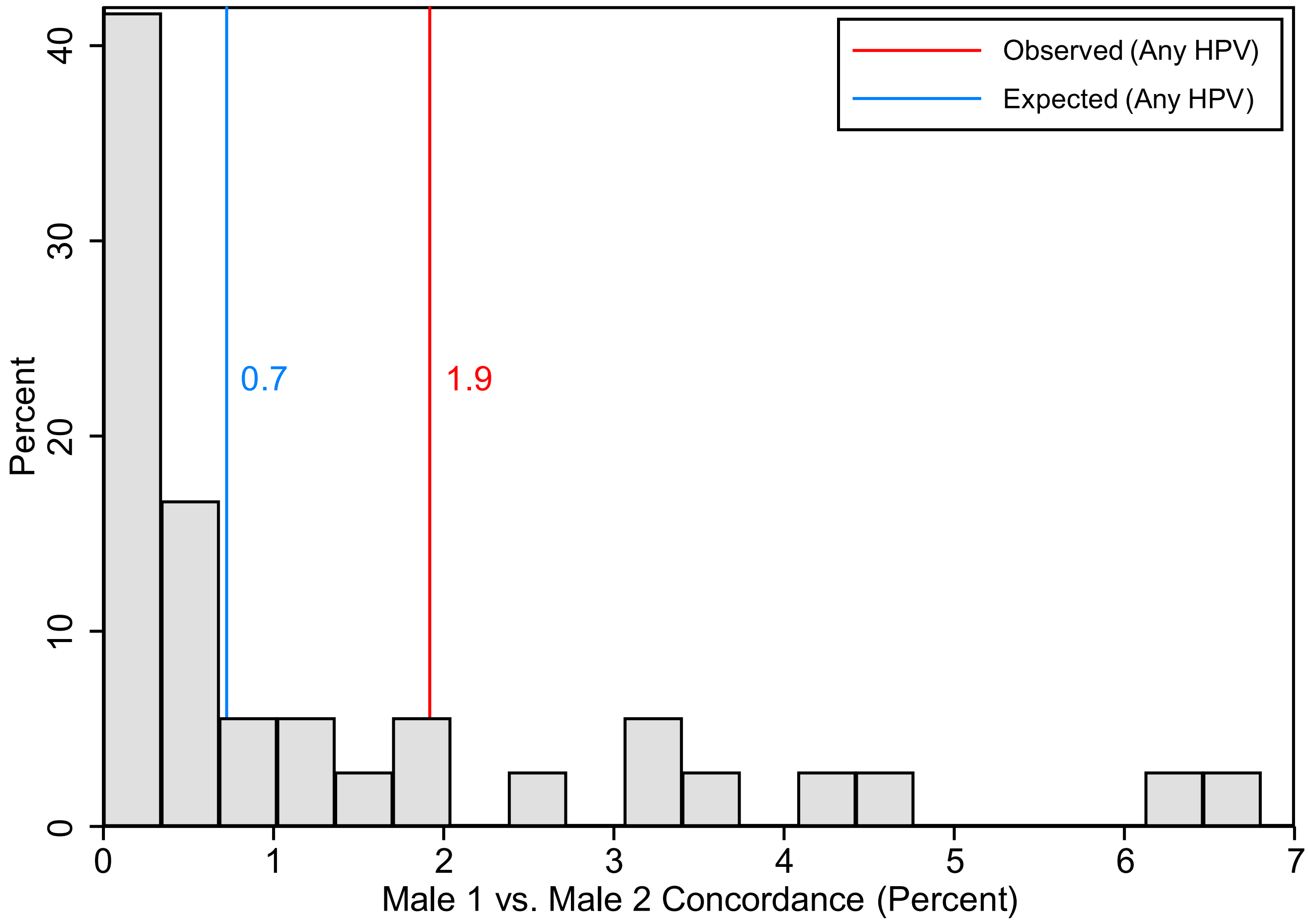


**Figure S2.** Type-specific expected human papillomavirus (HPV) concordance between males 1 and 2.

**Figure S2 Legend:** Distribution of expected concordance for 36 individual HPV types. Assuming the HPV positivity of males 1 and 2 is independent, the number of infections for which males 1 and 2 are expected to be concordant is equal to the product of the infection prevalence, divided by the total detectable infections (i.e., $\frac{P_{HPVx,Male1} \times P_{HPVx,Male2}}{T_{HPVx}}$).
